# Investigation of diffusion time dependence of apparent diffusion coefficient and intravoxel incoherent motion parameters in the human kidney

**DOI:** 10.1101/2024.03.03.24303581

**Authors:** Julia Stabinska, Thomas Andreas Thiel, Helge Jörn Zöllner, Thomas Benkert, Hans-Jörg Wittsack, Alexandra Ljimani

**Author notes:** Corresponding author: Thomas A. Thiel, Department of Diagnostic and Interventional Radiology, Medical Faculty, University Dusseldorf, Moorenstr. 5, 40225 Düsseldorf, Germany. These authors contributed equally (Julia Stabinska, Thomas Andreas Thiel).

## Abstract

**Purpose:** To characterize the diffusion time (Δ_eff_) dependence of apparent diffusion coefficient (ADC) and intravoxel incoherent motion (IVIM)-related parameters in the human kidney at 3T.

**Methods:** Sixteen healthy volunteers underwent an MRI examination at 3T including DWI at different Δ_eff_ ranging from 24.1 ms to 104.1 ms. The extended mono-exponential ADC, and IVIM models were fitted to the data for each Δ_eff_, and medullary and cortical ADC, (pseudo-) diffusion coefficients (D* and D) and flow-related signal fraction (f) were calculated.

**Results:** When all the data was used for fitting, a trend toward higher ADC with increasing Δ_eff_ was observed between 24.1 and 104.1 ms ([median and interquartile range]: 2.10 [1.94 2.26] to 2.36 [2.09 2.69] x10^−3^ mm^2^/s for cortex, and 2.24 [2.10 2.36] to 2.64 [2.49 2.78] x10^−3^ mm^2^/s for medulla). In contrast, no significant differences in ADC were found when only the data acquired at b-values higher than 200 s/mm^2^ was used for fitting. When the IVIM model was applied, cortical and medullary f increased significantly (cortex: 0.24 [0.22 0.30] to 0.54 [0.43 0.63] x10^−3^ mm^2^/s; medulla: 0.26 [0.23 0.38] to 0.50 [0.39 0.62] x10^−3^ mm^2^/s) and cortical D (cortex: 1.66 [1.59 1.71] to 1.24 [1.10 1.47] x10^−3^ mm^2^/s) and cortical D* (cortex: 9.10 [5.50 12.9] to 4.19 [3.84 5.73] x10^−3^ mm^2^/s) decreased significantly between 24.1 and 104.1 ms.

**Conclusion:** Renal perfusion and tubular flow substantially contribute to the observed increase in ADC over a wide range of Δ_eff_ between 24 and 104 ms.

## Introduction

Diffusion-weighted imaging is a versatile MRI technique that enables *in vivo* measurements of the translational motion of water molecules on the micrometer length scale, which is orders of magnitude lower than the nominal MR image resolution. During these random diffusion-driven displacements, water protons microscopically probe the tissue microstructure by interacting with cell membranes and macromolecules that impede their motion. Consequently, diffusion in biological tissues is considered restricted when water molecules are constrained within cells by impermeable barriers, or hindered when the motion of water molecules outside the cells is impeded but not completely confined by the presence of obstacles.

An important consequence of restricted and hindered diffusion in tissues is the dependence of DWI measurements on effective diffusion time (Δ_eff_), which is the time interval over which the water molecule displacements are sampled in a DWI experiment^1^. By adjusting the diffusion time, different diffusion length scales can be probed to provide sensitivity to unique properties of tissue microstructure. Overall, the presence of diffusion-restrictive barriers at relatively long diffusion times reduces the apparent diffusion coefficient^2^. Indeed, a significant decrease in diffusivity with increasing diffusion time was previously found in the brain^3^, breast^4^, liver^5^, and skeletal muscle^6,7^. Interestingly, we observed an opposite trend in the kidney where the DTI metrics (ADC and FA) increased with the diffusion time in a broad range of Δ_eff_ between 20 and 120 ms^8^. While the exact reasons for this are not yet fully understood, it is reasonable to assume that the time-dependent increase in renal diffusivity and anisotropy calculated using the mono-exponential tensor model can partially be attributable to strong vascular and tubular flow contributions to the overall DWI signal in the kidney.

It is well known from *in vivo* brain DWI that orientationally incoherent blood flow generates a “pseudo-diffusion” effect that can distort the measurements of mono-exponential ADC at relatively low b-values (<500 s/mm^2^)^9^. This effect, known as “intravoxel incoherent motion (IVIM)”, is especially pronounced in highly vascularized abdominal organs, such as the liver and the kidney. In normal kidneys, the estimated fractional vascular volume is around 25% - 40%^10^, which is much higher than that measured in the brain (approximately 5%). In addition to renal blood perfusion, tubular pre-urine flow and water reabsorption also enhance the IVIM effect in the kidney. Because the signal attenuation due to these flow-driven processes is much greater than attenuation caused by molecular diffusion, both contributions can be separated by using the IVIM model. The most commonly used model of the IVIM signals is the biexponential model, given by *S_b_* = *S*_0_ · ((1 − *f*) · *e*^−*b·d*^ + *f* · *e*^−*b·d*\*^), where *D* is the ‘true’ tissue diffusion coefficient and *f* and *D\** are the signal fraction and pseudo-diffusion coefficient of the microcirculatory compartment, respectively. Encouraging results using the IVIM model have been obtained in various renal pathologies, including allograft rejection^11^, renal masses,^12,13^ renal artery stenosis^14^ and renal dysfunction^15^. However, studies exploring the potential dependencies of the renal IVIM parameters on experimental pulse sequence timing, including diffusion time, are rare.

With the proper sampling and modelling, Δ-dependent DWI could be exploited to probe specific microstructural and vascular properties, such as the tubular diameter and geometry, membrane permeability and blood velocity. These parameters may be particularly sensitive to structural changes, such as tubular hypertrophy, renal interstitial fibrosis and tubular atrophy, which are associated with the progression of renal diseases. Finally, the Δ-dependency of IVIM signals could shed additional light into vascular and intratubular flow contributions to renal DWI. With this motivation in mind, we sought to characterize the diffusion time dependence of the IVIM-related parameters in the human kidney at 3T. In accordance with our previous work^8^ and the relatively large tubular diameters (20 to 50 μm^16^), we acquired IVIM data using a stimulated acquisition mode (STEAM) approach that allows the use of comparatively long diffusion times (beyond the T_2_-value of the tissue of interest) that are not feasible with conventional spin echo diffusion pulse sequence.

## Methods

### Healthy subjects

The study was approved by the local ethics committee, and written consent was obtained from each participant prior to MRI examination. Sixteen healthy subjects (7 females and 9 males; age range: 20–30 years; mean age: 24.9 ± 3.3 years) with no history of renal diseases were included in this study. Subjects were not given any restrictions regarding fluid or food intake.

### MRI data acquisition

MRI experiments were performed on a 3 Tesla MRI system (MAGNETOM Prisma, Siemens Healthineers, Erlangen, Germany) using an 18-channel torso array coil and a 32-channel spine coil. For anatomical visualization, three T2-weighted Half Fourier Single-shot Turbo spin-Echo (HASTE) images were acquired in a coronal-oblique plane through the long axis of both kidneys. DWI data was collected using a DWI 2D echo-planar imaging research application sequence with STEAM diffusion encoding and the following parameters: TR/TE = 1900/ 49 ms, FOV = 370 x 370 mm^2^, matrix = 176 x 176, Partial Fourier = 5/8, slice thickness = 5 mm, number of slices: 3, bandwidth = 1894 Hz/Px, GRAPPA factor = 2, fat suppression technique: SPAIR, 3D diagonal diffusion mode, 16 b-values: 0, 5, 10, 15, 20, 30, 50, 70, 100, 150, 200, 250, 300, 400, 500, 750 s/mm^2^, number of averages per b-value = 5. Diffusion gradient duration was fixed at 6.2 ms and effective diffusion times (Δ_eff_) were = 24.1, 44.1, 64.1, 84.1, and 104.1 ms with corresponding mixing times (‘*T1 relaxation period’*) of 9.8, 22.5, 42.5, 62.5, and 82.5 ms respectively. For the classical and STEAM diffusion encoding, b = γ²G²δ²(Δ_eff_ − δ_eff_/3). The DWI data acquisition was respiratory-triggered at the exhale phase of the respiratory cycle with a threshold of 20%. The average DWI acquisition time for each diffusion time was about 5 minutes and 30 seconds, depending on subject’s respiration rate.

### Data post-processing and analysis

Retrospective 2D registration based on a normalized mutual information similarity measure was performed to align all DW-weighted images at each b-value and diffusion time using an in-house developed software based on ANTs (http://stnava.github.io/ANTs/). Gaussian filter of 3 x 3 was then applied to the data, and a single slice that was least affected by the respiratory motion and contained clearly visible cortex and medulla was selected for further analysis. Cortex and medulla ROIs were manually segmented on b_0_ images by a research scientist (J.S. 7 years of experience) under the guidance of an abdominal radiologist (A.L. 10 years of experience) using ITK-SNAP (Version 3.8.0)^17^. The subsequent data analysis was performed using custom-written MATLAB routines (version R2021a, MathWorks, US). When performing experiments at different diffusion times with a STEAM acquisition, T_1_ decay needs to be taken into account as it varies with mixing time (TM)^18^. Figure 1 shows a simplified diagram of the STEAM-DWI sequence used in this study. The acquired STEAM-DWI signal was modeled using two different model functions: (1) *S_b_* = *S*_0_ · (1 − *e*^−*TR_eff_/T_1_*^) · *e*^−*TM/T_1_*^ · *e*^−*b·ADC*^ (*extended mono-exponential ADC model*), and (2) *S_b_* = *S*_0_ · (1 − *e*^−*TR_eff_/T_1_*^) · *e*^−*TM/T_1_*^ · ((1 − *f*) · *e*^−*b·D*^ + *e*^−*b·D*\*^) (*extended bi-exponential IVIM model*) with *TR_eff_* = *TR* − *TM* − 0.5 · *TE*, where TM is the mixing time. Mono-exponential ADC values at different Δ were estimated by least-squared curve fitting as described previously^19^. To enhance the stability of the estimated parameters (S_0_, ADC, and T_1_) all 16 b-values were initially used for fitting. In the next step, only a subset of the data acquired at b-values above 200 s/mm^2^ was fit to the same model in order to minimize the pseudo-diffusion signal contribution. In addition, T_1_ was fixed to the values determined in the previous step to reduce the number of free fitting parameters.

To improve the stability of the extended bi-exponential fitting, the Image Downsampling Expedited Adaptive Least-squares (IDEAL)^20^ approach was employed. First, the extended IVIM fitting of the DW signal obtained from the images downsampled to 1 x 1 with the loosely constrained lower and upper bounds was performed to determine initial values for the subsequent fitting step. These initial values and boundaries tightly constrained to 20% of the initial values for the signal fraction and relaxation times, 10% for the corresponding (pseudo-) diffusion coefficients, and 50% for the S_0_ images intensities were then used for voxel-wise fitting of the DW images downsampled to 2 x 2. The same steps were repeated for the images iteratively downsampled to 4 x 4, 8 x 8, 16 x 16, 32 x 32, 64 x 64, 96 x 96, 128 x 128, 152 x 152 until the original acquisition matrix size of 176 x 176 is reached. The initial parameter values for each voxel in each iterative step were determined by spatially interpolating the fitted parameters of the previous downsampled image using bilinear interpolation. The source code for the IDEAL fitting is freely available on GitHub (https://github.com/stabinska/IDEAL). All 16 b-values were used for the extended IVIM fitting to estimate D, D*, f, and T1.

**Figure 1:**
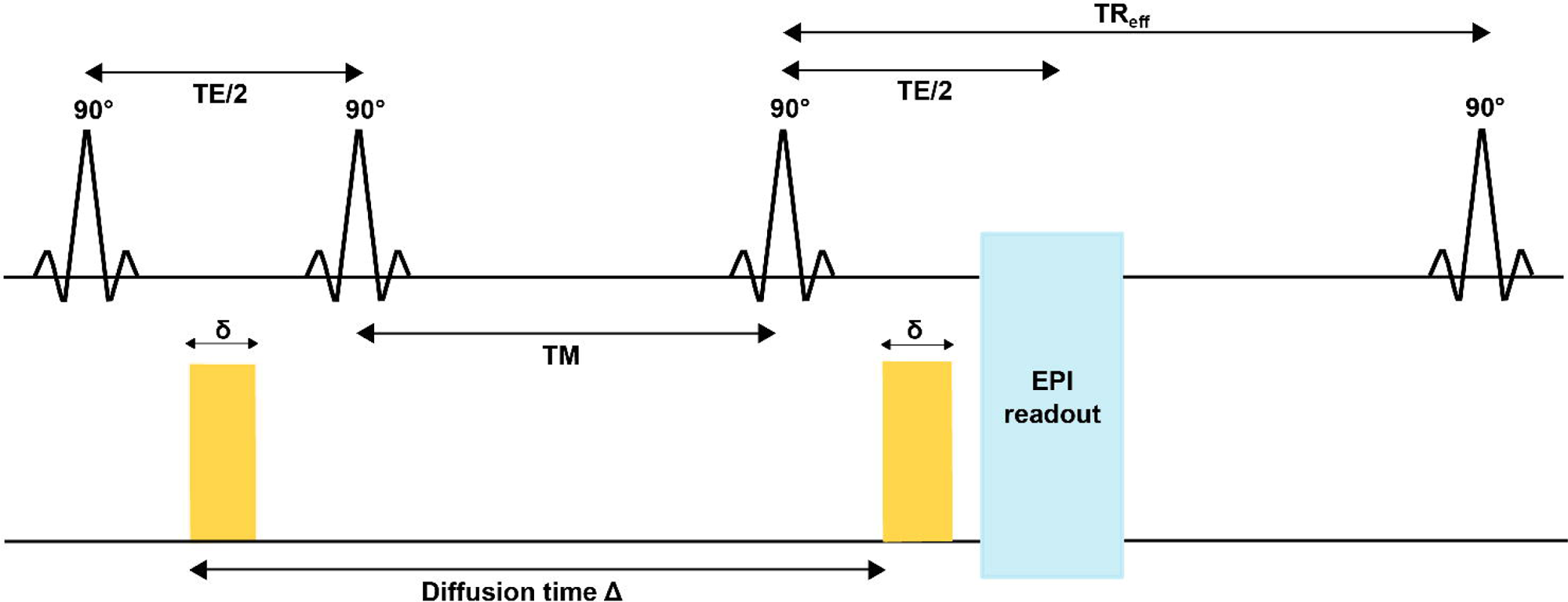
Simplified diagram of the STEAM pulse sequence used in this study. With three 90° pulses the signal of the stimulated echo is utilized for imaging. The diffusion gradient duration and TE were kept constant (δ = 6.2 ms, TE = 49 ms), while the diffusion gradient separation varied (= diffusion time, Δ) between acquisitions along with the mixing time (TM).

### Statistical analysis

Cortical and medullary ADC and IVIM parameter estimates obtained at different diffusion times were compared using ANOVA followed by a Bonferroni-corrected post hoc paired t-test. Statistical analysis was performed using an open-source R-package SpecVis (version 1.0.0) available on GitHub (https://github.com/HJZollner/SpecVis)^21^ and visualized using custom-written R-scripts (R-software, version 4.1.3, R Foundation For Statistical Computing, Vienna, Austria)^22^.

## Results

### Dependence of ADC on diffusion time

Figure 2 displays the cortical and medullary ADC obtained using all b-values data (Figure 2A) and a subset of the data acquired with b-values above 200 s/mm^2^ (Figure 1B). When all the data was used for fitting, ADC increased with diffusion time. In the cortex, ADC at Δ_eff_ = 44.1 ms (P = 0.016), Δ_eff_ = 84.1 ms (P = 0.016), and Δ_eff_ = 104.1 ms (P = 0.011) were significantly different from ADC at Δ_eff_ = 24.1 ms. In the medulla, ADC increased significantly between Δ_eff_ = 24.1 ms and Δ_eff_ = 84.1 ms (P = 0.006), between Δ_eff_ = 24.1 ms and Δ_eff_ = 104.1 ms (P = 0.001), between Δ_eff_ = 44.1 ms and Δ_eff_ = 84.1 ms (P = 0.004), between Δ_eff_ = 44.1 ms and Δ_eff_ = 104.1 ms (P = 0.0001), and between Δ_eff_ = 64.1 ms and Δ_eff_ = 104.1 ms (P = 0.009). In contrast, no significant differences in ADC were found when only the data acquired at b-values higher than 200 s/mm^2^ was used for fitting.

**Figure 2:**
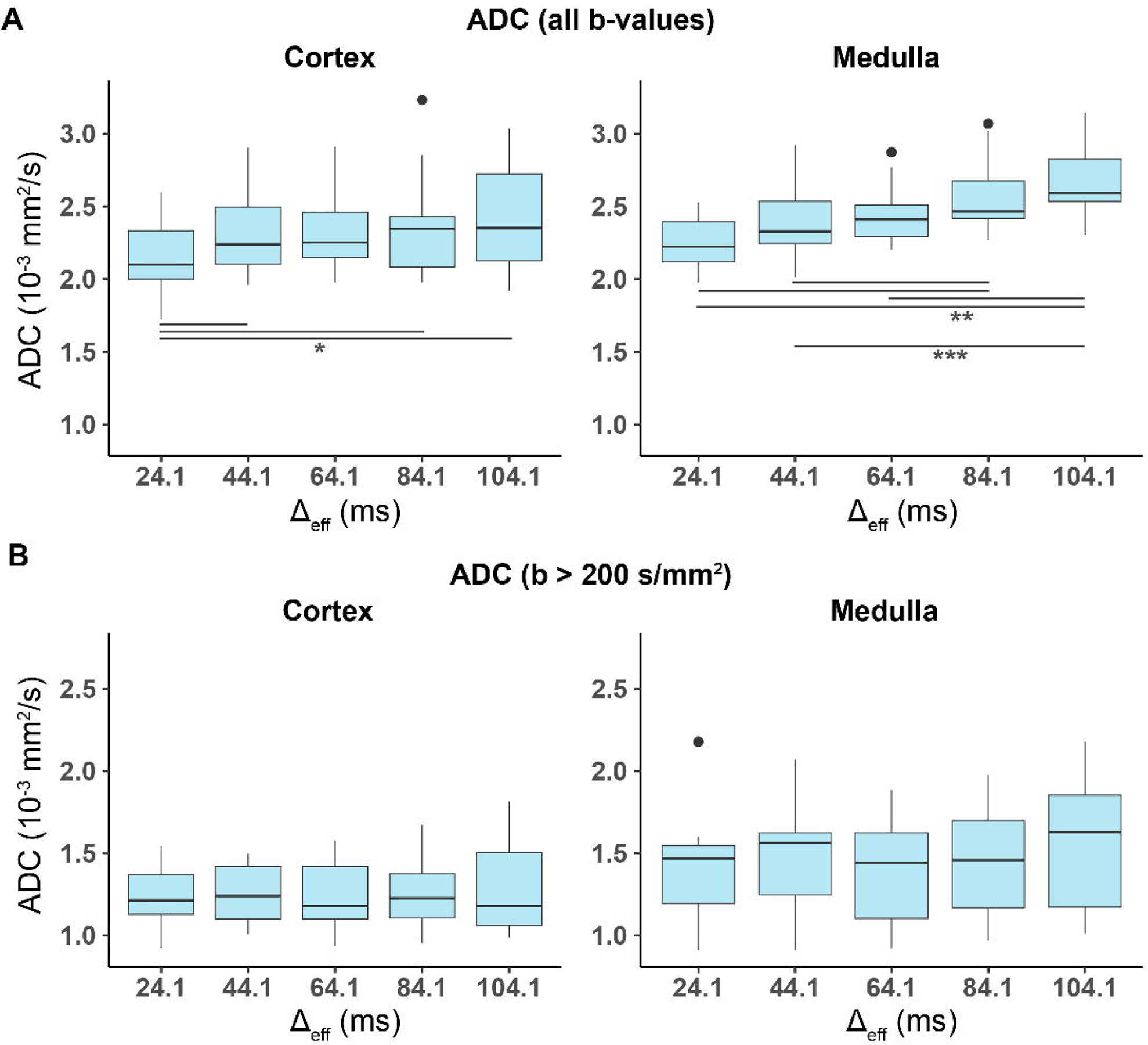
Distributions of the median values of the cortical and medullary ADC values obtained using data acquired (A) for all 16 b-values and (B) for b > 200 s/mm^2^ in all subjects. Asterisks indicate significant differences with P < 0.05 = *, P < 0.01 = **, and P < 0.001 = ***. ROI, region of interest.

### Dependence of IVIM parameters on diffusion time

Representative IVIM parameter maps obtained from the biexponential IVIM fitting at different diffusion times are displayed Figure 3. Overall, a trend toward a higher flow-related signal fraction and a lower diffusion coefficient and pseudo-diffusion coefficient with increasing diffusion time can be observed.

**Figure 3:**
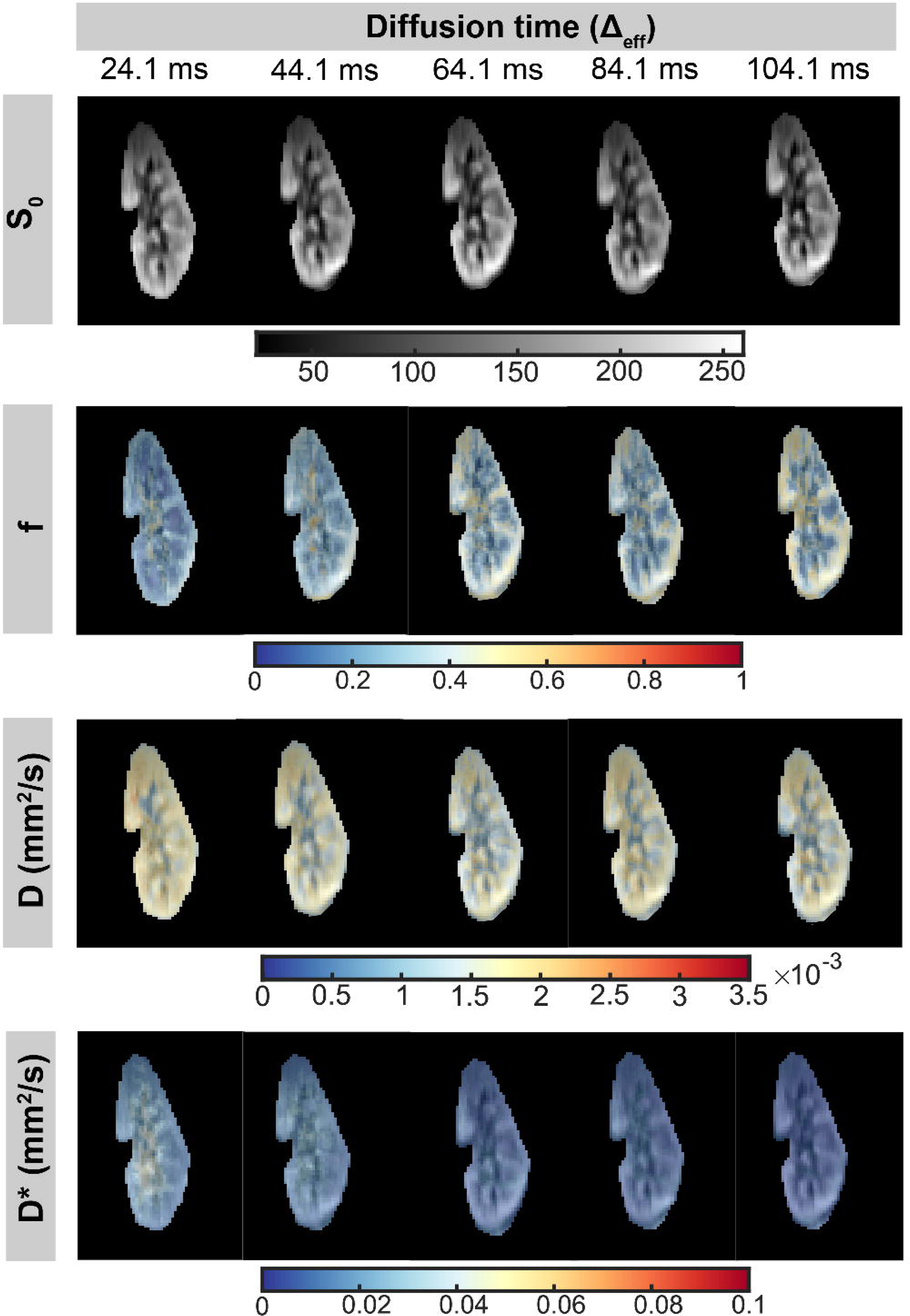
S_0_ image and IVIM parameters (f, D, and D*) maps obtained at different diffusion times using T1-corrected biexponential IVIM modelling.

Figure 4 shows violin plots of the IVIM parameters measured in the renal cortex and medulla across the investigated diffusion time range. Despite relatively large inter-subject variability, significant differences in f, D and D* obtained at different Δ were found. Cortical f increased significantly between Δ_eff_ = 24.1 ms and Δ_eff_ = 104.1 ms (P = 0.026). In the medulla, f at Δ_eff_ = 64.1 ms (P = 0.047), Δ_eff_ = 44.1 ms (P = 0.011) and Δ_eff_ = 24.1 ms (P = 0.011) were significantly different from f at Δ_eff_ = 104.1 ms, and f at Δ_eff_ = 44.1 ms (P = 0.017) and Δ_eff_ = 24.1 ms (P = 0.047) were significantly lower from f at Δ_eff_ = 84.1 ms. In contrast, diffusion and pseudo-diffusion coefficients showed a decrease with Δ. Cortical D at Δ_eff_ = 84.1 ms (P = 0.022) and Δ_eff_ = 104.1 ms (P = 0.009) were significantly different from cortical D at Δ_eff_ = 24.1 ms, while medullary D decreased significantly between Δ_eff_ = 24.1 ms and Δ_eff_ = 84.1 ms (P = 0.017). Significant differences in cortical D* were also found at Δ_eff_ = 64.1 ms (P = 0.017), Δ_eff_ = 44.1 ms (P = 0.017) and Δ_eff_ = 24.1 ms (P = 0.039) compared to cortical D* at Δ_eff_ = 104.1 ms. In the medulla, D* at Δ_eff_ = 84.1 ms (P = 0.039), Δ_eff_ = 64.1 ms (P = 0.017) and Δ_eff_ = 44.1 ms (P = 0.011) were significantly different from D* at Δ_eff_ = 104.1 ms. All fitting results are summarized in Table 1.

**Figure 4:**
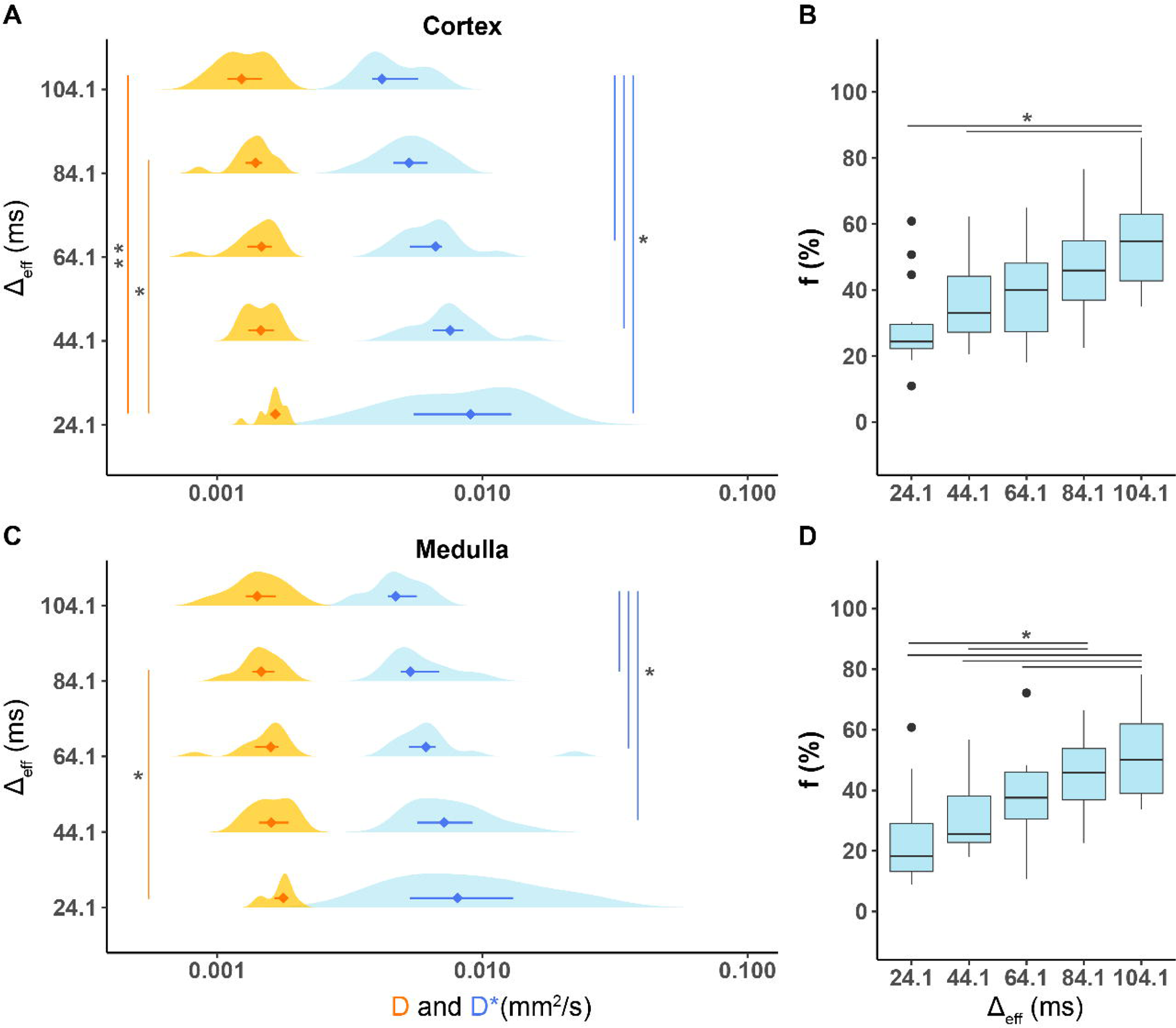
Distributions of the median values of the IVIM parameters (D, D*, and f) measured in the (A, B) cortical and (C, D) medullary ROIs in all subjects. Asterisks indicate significant differences with P < 0.05 = *, P < 0.01 = **, and P < 0.001 = ***. ROI, region of interest.

**Table 1:**
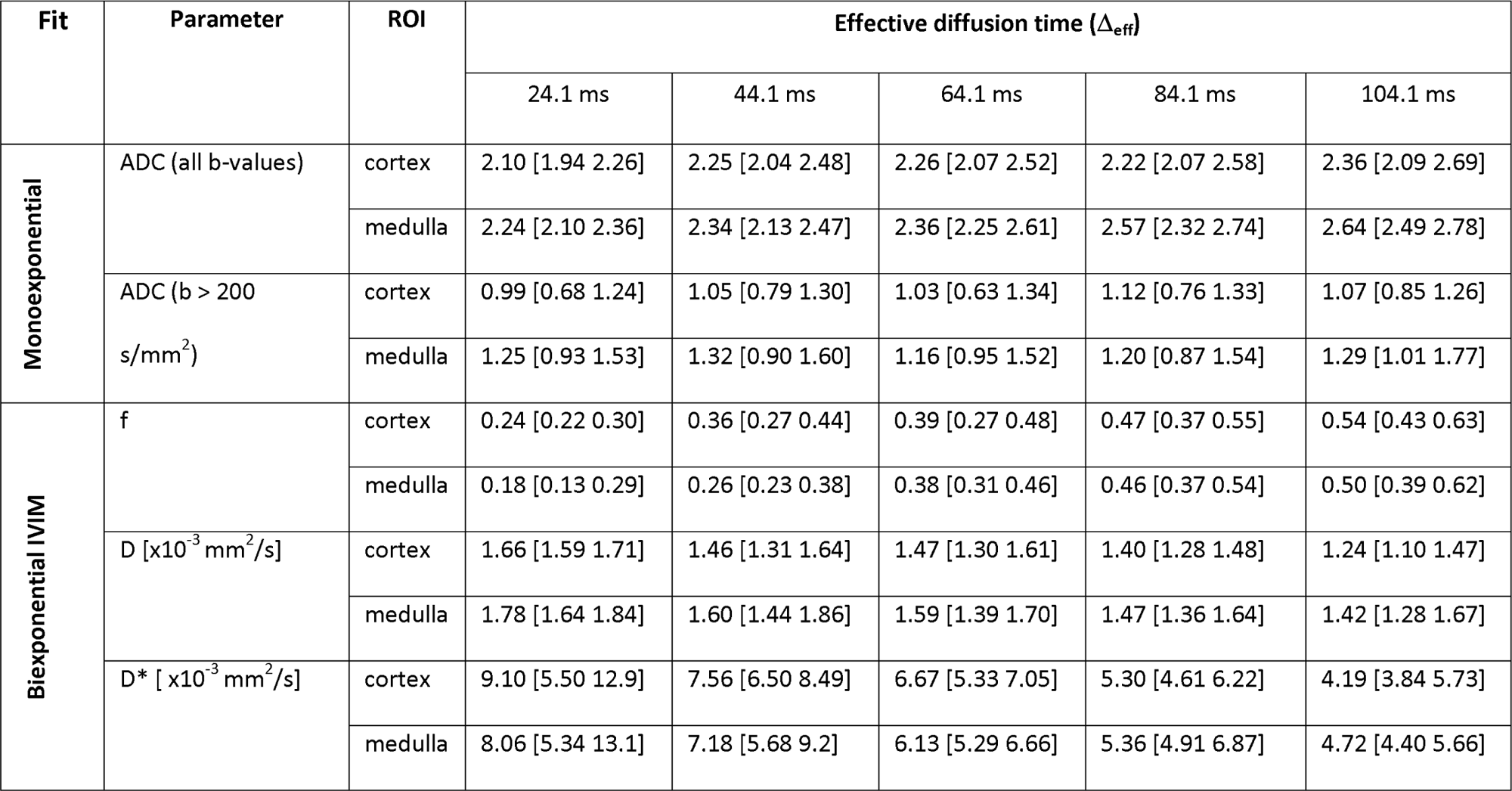
ADC and IVIM parameters obtained at different diffusion times. The values are presented as median, 25th, and 75th percentile.

## Discussion

In this study, we used a STEAM-DWI sequence to measure diffusion time-dependent changes in ADC and IVIM parameters in healthy human kidneys at 3T. In our previous work^8^, we demonstrated that renal diffusivity and fractional anisotropy obtained from the mono-exponential tensor model increase with diffusion time in a broad range of Δ_eff_ from 20 to 120 ms. We hypothesized that this trend might be partially due to strong vascular and tubular flow that contributes to the overall DWI signal measured in the kidney. Here, we investigated our hypothesis by applying a two-compartment IVIM model to renal DWI data acquired at multiple b-values.

The present study confirms our previous observations and provides experimental evidence that the signal fraction attributed to pseudo-diffusion increases significantly with diffusion time. Varying the diffusion time in STEAM acquisitions affects the T1-weighting, thus complicating the analysis of the diffusion time dependence. To partially overcome this limitation, we extended the conventional IVIM model used for fitting by an additional exponential term to account for T1 relaxation^18^. However, because the relaxation times of blood and tubular fluid - two main contributors to the IVIM signal at lower b-values (below 200 s/mm^2^) - differ substantially from the surrounding renal tissue, these two IVIM compartments will exhibit different T1-weighting at arbitrary diffusion times. Toward longer diffusion times, the signal attributable to moving blood and tubular fluid (compartment with fast diffusion and slow T1 relaxation) increases relative to the renal tissue signal (compartment with slow diffusion and fast T1 relaxation). As a result, the IVIM-derived signal fraction of the microcirculatory compartment in the kidney increases when measured with STEAM. Similar observations were made in the liver^23^ and pancreas^24^. However, in those studies both diffusion time and TE were varied and their influence on the estimated IVIM parameters cannot be separated. In order to only detect differences due to changes in Δ_eff_, TE was kept constant in our study.

When the mono-exponential DWI model is applied, the effect of the differences in T1 relaxation between blood/tubular fluid and tissue is reflected in the increasing ADC at longer diffusion times^25^. A potential solution to this issue is to incorporate the different T1 relaxation times into the IVIM model. However, introducing an additional free parameter in the fitting procedure results in less stable parameter estimations^26^. For more reliable analysis, extensive DWI data acquisition protocols would be required which entail longer acquisition times. More efficient T1-diffusion acquisition could be achieved with a novel STEAM-based technique, MESMERISED (for ‘Multiplexed Echo Shifted Multiband Excited and Recalled Imaging of STEAM Encoded Diffusion)^27^ that uses echo-shifting to exploit the long dead time in STEAM acquisitions. Another solution to separate the effects of T1 relaxation and diffusion time is to use a twice-refocused STEAM acquisition^25^, which ensures constant T1 weighting over a wide range of diffusion time.

In contrast to flow-related signal fraction f, a trend toward lower diffusion coefficient D and lower pseudo-diffusion coefficient D* at longer diffusion times was observed. While the decrease of the diffusion coefficients with increasing diffusion times is consistent with restricted diffusion, the water exchange between the tubular lumen and interstitial space during the mixing time is likely to affect the Δ_eff_ -dependency of the IVIM parameters. It is expected that for very long diffusion times, the diffusion coefficient will reach an asymptotic value rather than trending to zero. On the other hand, if Δ_eff_ is kept short enough so that the diffusion distance is smaller than the separation between the cell membranes, the diffusion will be largely unrestricted. Consequently, the effect of restricted diffusion can be modulated by adjusting the diffusion time. The diffusion time range probed in our study corresponds to the diffusion length scale of approximately 16 to 33 μm calculated using Einstein’s mean quadratic displacement equation <χ^2^>^1/2^ = (2ADC_0_ × t_diff_)^½^, where ADC_0_ the ADC limit at b_0_ defined as ADC_0_ = f·D + (1 - f) ·D* (the averaged ADC_0_ in our study was 5.17 x 10^−5^ mm^2^s^−1^)^28^. Note that, as the tubular luminal diameters vary between 20 um and 50 μm in the human kidney, the diffusion time range selected in this work enables sensitivity to the length scale of a relatively wide range of tubular segments (from the thin segments of the loop of Henle to the proximal and distal convoluted tubules). Longer diffusion times could be achieved with STEAM to further extend the diffusion length scale. However, the relatively low SNR of the STEAM DWI sequence, which precludes accurate estimation of the (pseudo-) diffusion coefficients^29^, could be a limiting factor.

Limitations of this study include the lack of a gold standard to estimate the actual vascular and tubular fractions that appear to strongly vary across the subjects, and the limited range of diffusion times due to hardware constraints and SNR issues at higher diffusion times. Further, the two-compartment IVIM model used here only considers a single case of fast microcirculatory flow and does not distinguish between the vascular perfusion and tubular pre-urine flow. Recent studies, however, demonstrated that the intra-tubular compartment introduces an intermediate-decay component in the renal DWI signal^30,31^ and that the three-compartment IVIM model better reflects the complex characteristics of renal tissue.

Despite the relatively small study population and above-mentioned limitations, our experimental findings shed light on the diffusion time-dependent nature of DWI/IVIM estimation in the kidney. To the best of our knowledge, this is the first study in the kidney providing initial evidence that diffusion time is a relevant acquisition parameter to be considered when interpreting and comparing IVIM imaging results from different studies. Controlling diffusion time should allow IVIM to achieve greater reproducibility between different sites and vendors. The next logical step for future investigations of the diffusion time dependence of renal diffusion is to develop an extended IVIM model that takes into account potentially confounding factors such as the differences in the relaxation times of each compartment and water exchange between the tubular lumen and medullary interstitium.

## Conclusion

In conclusion, our experimental results provide insights into the diffusion time dependent diffusion behavior in the kidney, indicating that renal perfusion and tubular flow substantially contribute to the observed increase in ADC over a wide range of diffusion times between 24 and 104 ms.

## Data Availability

All data produced in the present study are available upon reasonable request to the authors.

## Acknowledgement

The study was supported by a grant from the German Research Foundation (DFG) (project number: 408765040). Alexandra Ljimani is supported by an internal research grant of the local Research Committee of the Medical Faculty of Heinrich-Heine-University Düsseldorf (2020-65). Open Access funding enabled and organized by Projekt DEAL.

